# A new liability-based rare-variant burden test identifies a novel genetic association between *HTRA3* and idiopathic pulmonary fibrosis

**DOI:** 10.64898/2026.11.17.26368326

**Authors:** Sung Chun, Ahmad Samiei, Lauren Flynn, Heidi Makrynioti, Matthew Moll, Anna L. Peljto, David A. Schwartz, Michael H Cho, Shamil R Sunyaev, Ivan O Rosas, Gary M. Hunninghake, Benjamin A Raby

## Abstract

In contrast to large-effect monogenic variants, which are highly penetrant regardless of genetic background, variants with more modest effect sizes depend on a carrier’s genetic background to manifest disease. For many complex traits, including idiopathic pulmonary fibrosis (IPF), polygenic risk scores (PRS) have been shown to capture a significant proportion of the genetic background risk. While a rare-variant association test incorporating a PRS covariate to account for the genetic background has been demonstrated to improve statistical power for gene discovery in quantitative traits, for binary traits, this approach shows limited or no power gain over PRS-unadjusted tests. Here, we propose a novel rare-variant association test for binary traits that jointly models the contribution of rare variants and PRS under a liability threshold model. In simulated datasets, our new method, liability threshold burden test (LTBT), shows a well-controlled false positive rate and outperforms the sensitivity of existing methods. To demonstrate this new method, we apply it to sequencing datasets for IPF, a rare but highly morbid lung disease. By accounting for an individual’s genetic background with PRS, we identify and formally replicate a significant novel association of rare predicted damaging missense variants in *HTRA3* (HtrA serine peptidase 3) with IPF.

## INTRODUCTION

Rare variant association studies (RVAS) aim to map disease genes by identifying differences in the distribution of rare variants between cases and controls within target genomic regions.^1–3^ Historically, RVAS have been applied primarily to protein-coding regions; ^4–8^ however, recent expansions in large-scale biobanks providing whole-genome sequencing (WGS) data have facilitated their application to non-coding regulatory regions. ^9,10^ Because association signals are inherently faint for individual rare variants, RVAS boost statistical power by collapsing the signals across all rare variants within a given target region. To date, two statistical frameworks have emerged for RVAS: burden tests and overdispersion tests. Rare-variant burden tests are designed to detect the overall excess or depletion of causal rare variants in affected individuals under the assumption that they share a fixed effect size.^11–13^ In contrast, overdispersion tests allow the effect size of causal variants to be a random variable with non-zero variance.^14^ Neither approach, however, explicitly accounts for the interaction of rare variants with the carrier’s genetic background.

In many monogenic disorders, pathogenic mutations confer differing disease risks depending on genetic modifiers in the background, frequently leading to incomplete penetrance.^15–19^ Although most genetic modifiers cannot be identified individually with typical sample sizes, a significant proportion of their aggregate effect can be captured in the form of polygenic risk scores (PRS).^20–24^ In the past, PRS had been reserved for common disorders since large-scale genetic data required for PRS training are not readily available for rare diseases. For some disorders, however, rare monogenic variants coexist with a more common polygenic risk background in the general population. In such cases, a PRS can be constructed using genetic data from a genome-wide association study (GWAS) of the common complex form to capture the aggregate effect of genetic modifiers.^17,25^ A more recent study suggests that the interactions between rare monogenic variants and PRS of common phenotypes are more extensive than previously thought,^26^ which demonstrates that accounting for the joint effect of rare variants and PRS is not only feasible but also highly relevant.

Thus, in principle, a joint model incorporating both rare variants and PRS should boost the statistical power of RVAS by reducing the unexplained variance in the model. Indeed, a simple yet powerful strategy to include a PRS as a covariate into current RVAS methods demonstrated that this approach can significantly increase statistical power for RVAS in quantitative traits.^27^ For binary traits, however, the inclusion of a PRS covariate did not substantially improve statistical power. Unlike with quantitative traits, the threshold effect for phenotypic manifestation in binary traits may make rare variants interact nonlinearly with a PRS. As such, additive modeling of these factors may not be sufficiently effective to account for the genetic background in binary trait RVAS.

Previously, several studies have proposed to study relationships between rare variants and a PRS in case-control cohorts using a liability threshold model.^28–30^ As originally developed to explain the threshold behavior of binary traits,^31^ the liability threshold model can provide an ideal statistical framework to integrate the burden of rare variants and the genetic background captured by a PRS. Here, we introduce a new RVAS test that explicitly parameterizes the interaction between rare variants and a PRS under a liability threshold model. In simulations, we show that the new approach yields higher statistical power for disease gene discovery than current rare-variant association tests for binary traits, regardless of whether a PRS is incorporated as a covariate.

To demonstrate the utility of our new method, we apply it to idiopathic pulmonary fibrosis (IPF), a chronic late-onset lung disease characterized by progressive, irreversible interstitial fibrosis.^32,33^ Although the largest rare-variant association study to date for IPF (2,180 cases and 2,457 controls) replicated significant associations of two known genes (*TERT* and *RTEL1*) but it did not uncover any novel genes, leaving open the question about whether additional rare-variant associations can be identified.^34^

Since IPF is a complex trait, GWAS have also been conducted, successfully uncovering 14 genome-wide significant loci associated with the disease.^35^ The 14-SNP PRS constructed from this dataset achieves one of the highest prediction accuracies reported to date (explaining 18% of disease liability at ages 65+),^36^ rendering IPF a model phenotype for demonstrating the statistical power of our new method. By applying this method to sequencing data for IPF, we identify and formally replicate a novel rare-variant association in *HTRA3* (HtrA serine peptidase 3) with IPF.

## RESULTS

We propose a new rare-variant association test that explicitly models the interaction between rare variants and PRS for a binary trait (**Figure 1A**). Our approach, called a liability threshold burden test (LTBT), attempts to associate the probability of having disease with the hidden total liability underlying disease using a liability threshold model (**Methods**). We define the total liability as the additive sum of rare-variant effect (*γ*), PRS, and unexplained risk factors. Under a liability threshold model, the disease outcome is determined by whether an individual’s total liability is over the threshold determined by the prevalence of disease in population. If a rare variant confers a substantially larger effect compared to the liability threshold, the penetrance becomes nearly complete for its carriers (**Figure 1B**). This is because the rest of liability components (PRS and unexplained factors) is unlikely to be protective enough to counteract the rare variant by random chance. In contrast, if a rare variant has a modest effect, the chance of disease manifestation becomes highly dependent on the presence of other risk factors in the carrier. For instance, for a modestly pathogenic variant with *γ* = 1.0 (corresponding to the odds ratio of 12.2; **Figure 1C**), the penetrance will be low (5.8%) although this is still high compared to the simulated population prevalence (0.5%).

**Figure 1.**
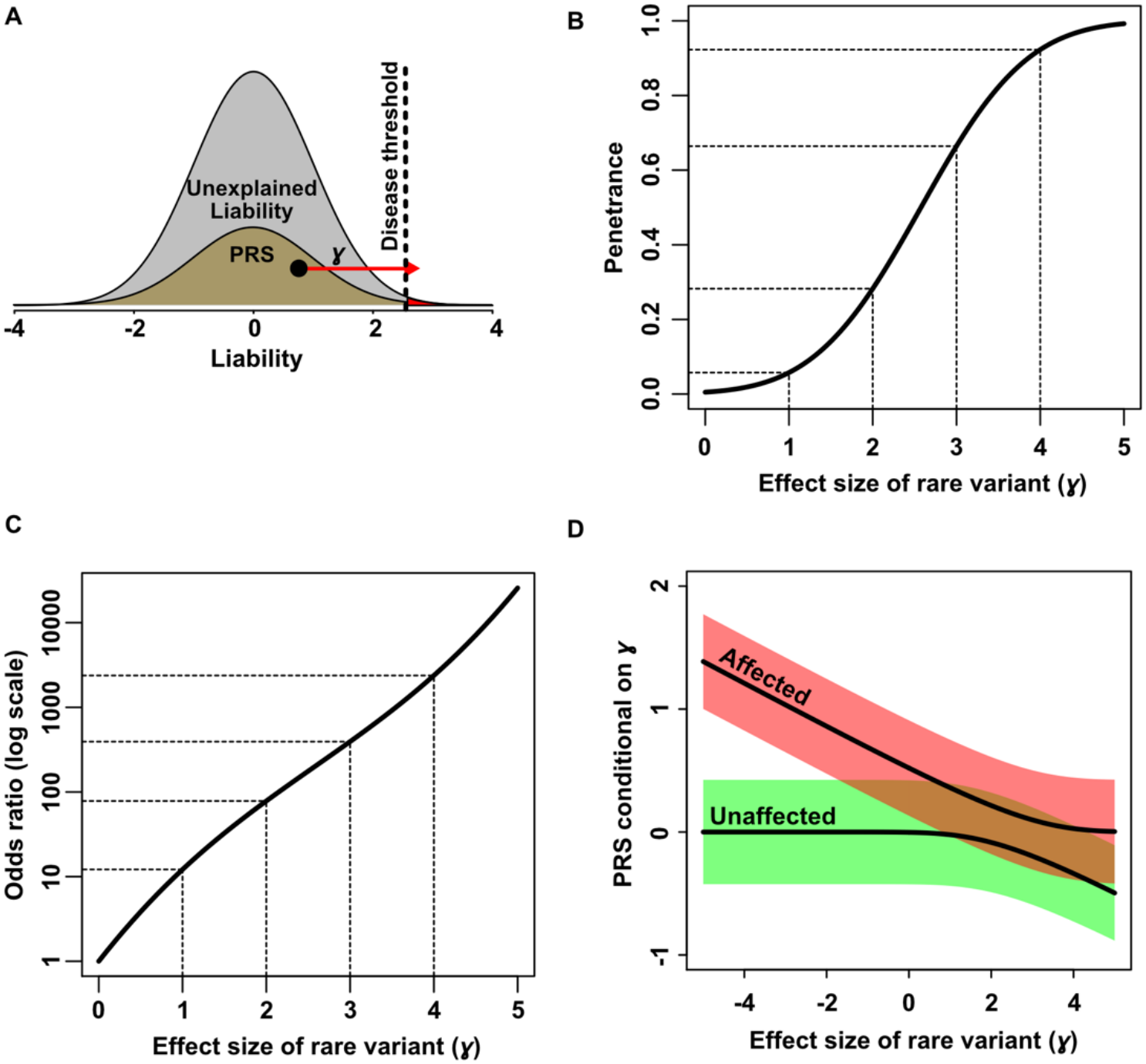
Statistical genetic rationale behind liability threshold burden test (LTBT). **(A)** Under a liability threshold model, the total liability of an individual is the sum of rare-variant effect (*γ*, red arrow), polygenic risk score (PRS, brown), and unexplained factors (grey). The disease outcome is determined by whether the total liability exceeds the disease threshold, determined by the prevalence. **(B)** The penetrance of rare variants is modest when *γ* is relatively smaller compared to the liability threshold. **(C)** The relationship between odds ratios *γ*. **(D)** The conditional distribution of PRS given *γ* among cases (red) and controls (green). The shaded areas indicate +/-SE around the conditional mean PRS. Simulation parameters: Prevalence = 0.5%. PRS explains 18% of heritability.

Conversely, if we already know the effect size of a rare pathogenic variant, we can infer the conditional distribution of the rest of the liability components, particularly of PRS, among carriers in affected and healthy subjects (**Figure 1D**). For example, for an affected carrier, the conditional mean of PRS is high if the rare variant has a protective or mild risk effect. In comparison, the conditional mean approaches 0 as the effect size of a rare variant becomes larger (i.e., rare variants with large enough *γ* are highly penetrant irrespectively of genetic background). Similarly, the conditional mean PRS of healthy controls is 0 if the variant has a protective or no effect, whereas it decreases toward the negative direction (protective background) as the rare variant has a larger *γ*. The degree to which the conditional distribution of PRS responds to the effect size of a rare variant in carriers is determined by the predictive power of PRS, i.e., how much of the liability is explained by PRS (denoted by 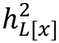). 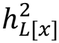 can be inferred from the predictive power of PRS, measured in the Area Under the Curve (AUC), in a case-control cohort.^37^

Using the conditional distribution of PRS given the rare variant effect size *γ*, we compare the likelihood of observed data under the null (*γ* = 0) and alternative (*γ* ≠ 0) hypotheses. The observed data consists of the disease outcome, genotypes of rare variants, and PRS for each individual. For computational efficiency, we assume that the heritability contributed by rare variants of each gene is negligible. We further assume that all rare variants have an equal fixed effect size and act dominantly to each other. Here, we approximate the conditional PRS with a normal distribution similarly as in Wray et al.^37^ In simulation, the normal approximation closely follows empirical conditional distributions of PRS across a wide range of simulated rare-variant effect sizes (**Supplementary Figure 1**). The p-value of rare-variant association test is calculated using the *χ*^2^-distribution with one degree of freedom under the nested log likelihood ratio test framework.

To test if our proposed approach achieves a well-controlled false positive rate, we generated 10^7^ simulated case-control cohorts under the null hypothesis (**Table 1**). For the simulated dataset, we sampled PRS and unexplained liability from a normal distribution under 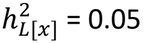, 0.1, and 0.2, covering typical predictive power of PRS. For rare variants, we randomly sampled their genotypes from the UK Biobank Whole-Exome Sequencing (WES) data, using the empirical distribution of predicted loss-of-function (pLoF) variants with minor allele frequency (MAF) < 1%. The cumulative allele frequency of all rare pLoF variants in a given gene was controlled to be in the range of 0.1% to 0.5%. Finally, we generated case-control cohorts of various ratios (1:1, 1:5 and 1:10) by down-sampling from a simulated unascertained population with the assumed disease prevalence of 0.5, 1, and 5%. In these simulated case-control cohorts, we let LTBT estimate 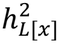 of PRS directly from the provided cohort but assume the prevalence of disease is known from larger external epidemiological data. Across all simulated parameters, we observe that the false positive rates of LTBT are well-controlled at significance levels (*α*) of 10^−3^, 10^−4^, and 10^−5^.

Next, we compared the sensitivity of our method with that of existing rare-variant association tests at the *α* level of 10^−6^ using simulated benchmark datasets. For competing methods, we considered a logistic regression model, Fisher’s Exact Test, and Sequence Kernel Association Test (SKAT)-O.^38^ Recently, it has been proposed that a simple strategy of adding PRS as a covariate to existing burden tests can improve statistical power of rare-variant association test for quantitative traits.^27^ To examine the advantage of our parametric approach against such non-parametric adjustment schemes in binary traits, we also compared our method with existing rare-variant association tests adjusting for PRS as a covariate. For the simulated benchmarks, we generated 1,000,000 balanced case-control cohorts of 5,000 cases and 5,000 controls for each simulation parameter regime by using the same simulation procedure as the null simulation. The effect size of pathogenic rare variant was varied from 0.2 to 1.0 on the liability scale, and the heritability explained by PRS 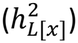 increases from 0 to 0.2 simulating the increasing accuracy of PRS. The advantage of LTBT was highest when the effect size of a rare variant was modest (*γ* = 0.4) in these simulation conditions (**Figure 2**). The statistical power of LTBT increases with higher PRS accuracy. For example, for a PRS with 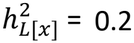, LTBT showed 18.1% higher statistical power (41.4%) to detect the burden of rare variants with *γ* = 0.4 compared to the second-best method, SKAT-O without a PRS covariate adjustment (35.0%). The advantage of non-parametric adjustment with a PRS covariate was not clear for binary traits in our simulations, as previously reported.^27^ Since PRS is correlated with the presence of rare pathogenic variants due to the thresholding effect in binary traits (**Figure 1D**), the statistical advantage of PRS covariate adjustment observed in quantitative traits do not necessarily translate for binary traits. LTBT showed consistently higher statistical power than other methods when we explored different disease prevalence in our simulation (**Supplementary Figure 2**). For a modestly predictive PRS with 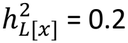, LTBT showed 10.4, 13.0, and 9.1% better power to detect the burden of rare variants with *γ* = 0.4 compared to the second-best method (SKAT-O) at the simulated disease prevalence of 0.5, 1, and 5%, respectively. Interestingly, when the prevalence is 5%, both logistic regression and SKAT-O start to show consistent improvements in statistical power when adjusting for a PRS covariate, although LTBT still outperformed these approaches (**Supplementary Figure 2C**).

**Figure 2.**
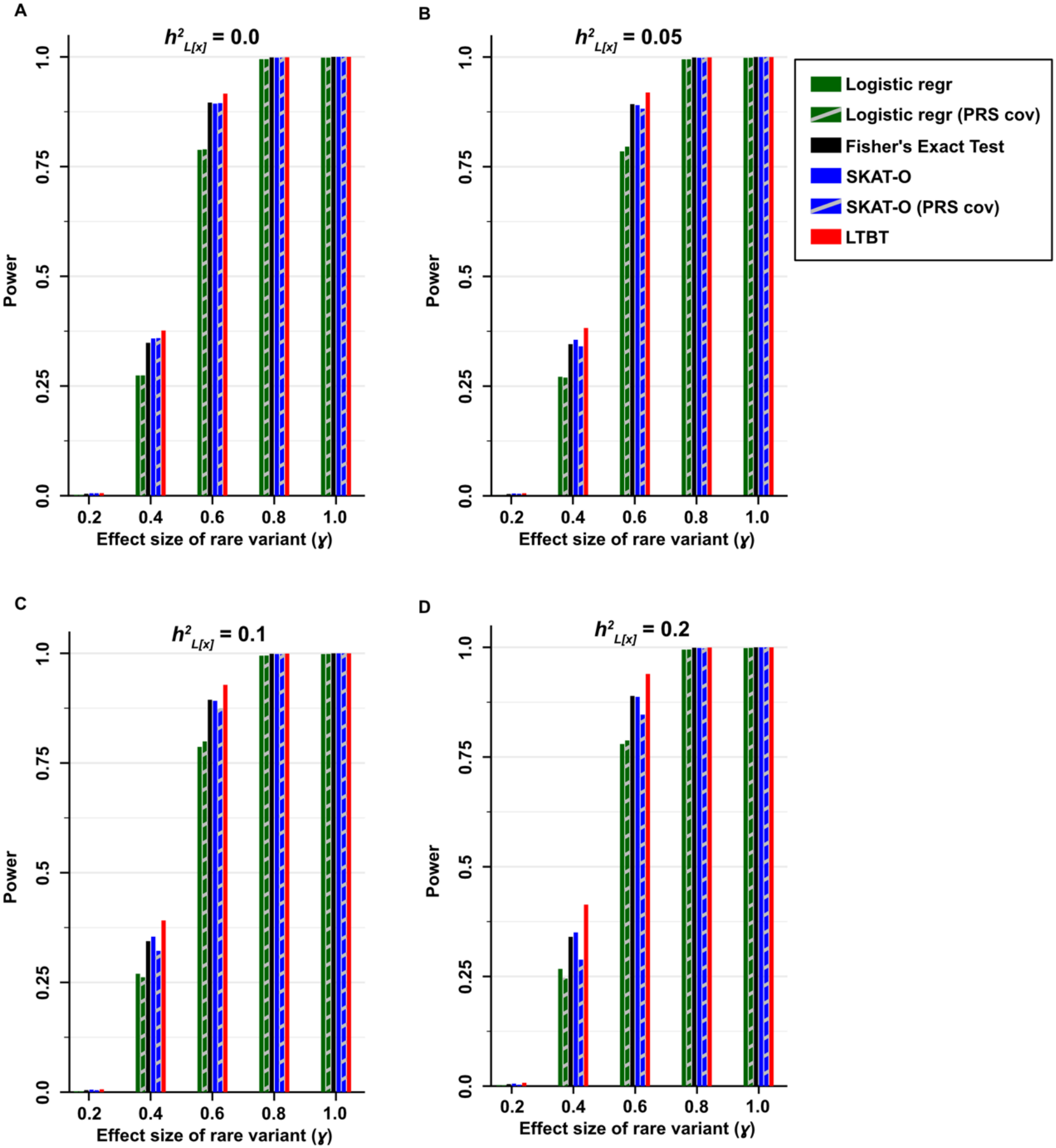
Statistical Power of Liability Threshold Burden Test (LTBT) and existing methods. LTBT and existing methods were compared in benchmark datasets simulated with PRS accuracies of 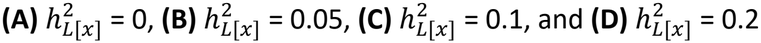. For each parameter, 1,000,000 case-control cohorts of 5,000 cases and 5,000 controls were generated. Prevalence = 0.005.

To demonstrate the advantage of our method in real data, we applied LTBT to WES data for IPF. The prevalence of IPF at the age of 65 or higher is estimated to be ~0.5% based on a large epidemiological study.^39^ For sequencing data, we used 136 IPF patients of European ancestry from the CGS-PF cohort ^40^ with 230,044 matched healthy controls from the UK Biobank.^6,8^ The IPF PRS constructed with the 14 genome-wide significant SNPs^41^ showed high prediction accuracy 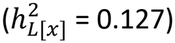 in this cohort although it is slightly lower than previously reported.^36^ To minimize biases due to the differences in data processing between cases and controls, we applied the same variant calling and quality control (QC) pipeline to cases and controls. Overall, we did not observe any genomic inflation in our LTBT statistics which may indicate technical bias or stratification between cases and controls (*λ*_*GC*_ = 0.89 for probably damaging missense variant scan; **Supplementary Figure 3**). We applied our test to two functional categories of rare variants with MAF < 1%, namely those annotated with pLoF by LOFTEE^42^ and those predicted to cause probably damaging missense mutations by PolyPhen.^43^ Three previously known IPF genes (*RTEL1, PARN*, and *TERT*) were replicated in our discovery cohort by both Fisher’s Exact Test and LTBT (P < 0.05, **Table 2**). The estimated average effect sizes for rare variants in these genes are in the range of 1 ≤ *γ* ≤ 2, which indicates that their penetrance in general population is modest (**Figure 1B**). Interestingly, LTBT estimates that probably damaging missense variants in *TERT* are slightly more pathogenic (*γ* = 1.60) than predicted loss of function variants in *RTEL1* (*γ* = 1.46). This estimation by LTBT was driven by the observation that affected carriers of rare probably damaging missense variants in *TERT* tend to have low PRS values (**Figure 3**). By taking advantage of such a signal, LTBT estimates a more significant p-value (P = 5.4 × 10^−5^) than by odds ratios alone (Fisher’s Exact Test P = 4.1 × 10^−3^).

**Figure 3.**
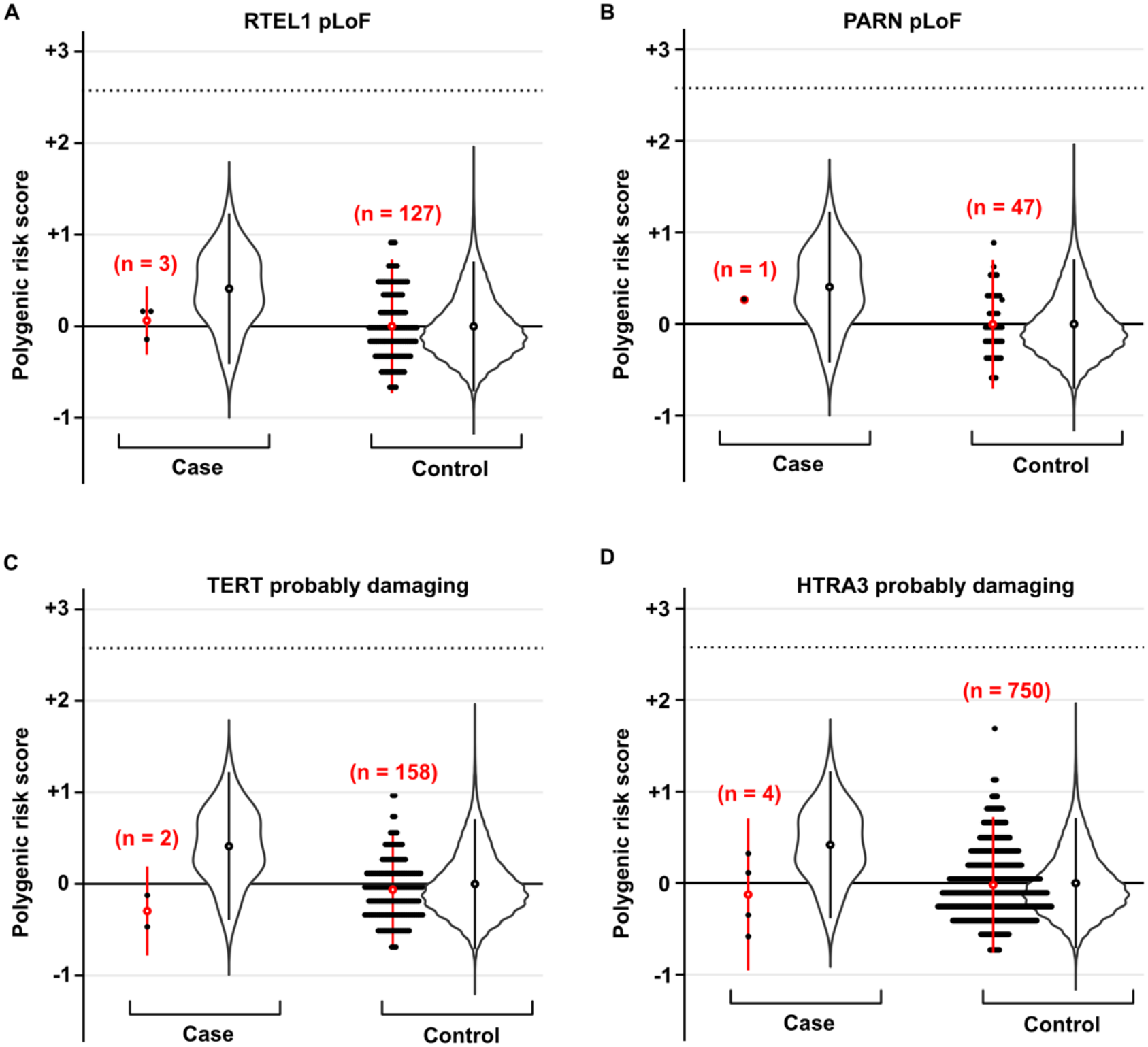
Distribution of polygenic risk scores in cases and controls. Each solid dot represents carriers of rare variants. Violin plot indicates the distribution of non-carriers. Open circle denotes the average. Vertical lines around open circles represent standard error.

To discover novel genes, we expand our analysis to all genes with cumulative allele frequency of rare variants to be at least 0.1% in our discovery cohort, for which we have strong statistical power. A total of 219 and 5,127 genes satisfy this criterion for pLoF and probably damaging missense rare variants, respectively. For the pLoF, no gene passed the false discovery rate (FDR) threshold at 0.05. For the probably damaging missense variants, a total of four genes, including *HTRA3* (HtrA serine peptidase 3), had FDR < 0.05 (**Supplementary Table 1**).

Similar to known genes, the PRS of affected carriers of rare variants in *HTRA3* were shifted lower than that of affected non-carriers (**Figure 3D**). Reflecting this, the p-value of LTBT (P = 1.8 × 10^−4^) was more significant than that of Fisher’s Exact Test (P = 1.1 × 10^−3^).

Four candidate genes satisfying FDR < 0.05 in the discovery cohort were moved forward to formal replication in All of Us biobank.^44^ Since the prediction accuracy of PRS declines outside of the original GWAS population (European ancestry), we conducted the validation analysis in a subset of participants with matching ancestry. To identify IPF cases with the highest confidence, we combined the diagnosis of IPF in medical records with the history of anti-fibrotic medication use. In total, we obtained 90 IPF cases and 106,831 controls with the ages 65+. Of the four candidate genes, *HTRA3* was formally replicated after Bonferroni correction (Fisher’s Exact Test P = 6.5 × 10^−3^ and LTBT P = 0.012, **Table 2**).

## DISCUSSION

In the current study, we present a new rare-variant association test that integrates the burden of rare variants with their genetic background captured by PRS. The new method provides significantly higher statistical power than existing methods in simulation. A recent study proposed to adjust rare-variant association tests by incorporating a PRS covariate to account for the genetic background effect.^27^ However, it has not been clear why the covariate adjustment could not improve statistical power for binary traits. Here, we show that to account for the genetic background in binary traits, we need to explicitly model the burden of rare variants under a formal liability threshold model. This is because the thresholding effect of disease manifestation causes the conditional dependency between the burden of rare variants and PRS in cases and controls. As such, simple strategy of including a PRS covariate into existing models cannot consistently boost statistical power of rare-variant association test for binary traits.

By applying our new method to IPF, we identified a novel association of rare probably damaging missense variants in *HTRA3* and formally replicated the signal in an independent validation cohort. A recent study of cardiac fibroblasts implicated this gene in the pathogenesis of cardiac fibrosis via TGF-β signaling.^45^ Moreover, the knockout of *HTRA3* was shown to affect epithelial-mesenchymal transition (EMT) in bleomycin-induced pulmonary fibrosis model in a mouse.^46^

Recently, the lack of novel rare-variant burden signal in Trans-Omics for Precision Medicine (TOPMed) IPF study has sparked a debate on whether more IPF genes can be identified.^34^ Our study suggests that more novel IPF genes such as *HTRA3* remain to be found. To improve statistical power for the discovery of disease genes driven by lower penetrance rare variants, we need a statistical test that integrates the rare-variant burden with the genetic background. While our proposed method addresses part of this, there remains limitations in the current approach. The current method does not allow for the variation of effect sizes as in overdispersion-based tests. Also, it assumes that the disease prevalence is known from a large epidemiological cohort and that the case subjects can be identified by applying the same diagnostic criteria. Furthermore, since our method assumes a uniform prediction accuracy of PRS, it cannot be applied to mixed-ancestry population in the current form. Future methodological studies are warranted to address these limitations for rare polygenic disorders including IPF.

## METHODS

### Statistical model

Our method, called liability threshold burden test (LTBT), is based on the following genetic model:

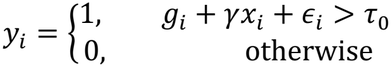

where *y*_*i*_ is the disease outcome (1 for diseased and 0 for healthy), *g*_*i*_ is the PRS, *x*_*i*_ is the genotype of rare variants (1 for a carrier and 0 for a non-carrier under a dominant model), and *ε*_*i*_ is the unexplained liability for each individual *i*. The genotype-phenotype relationship of this model is specified by two parameters: *γ* representing for the effect size of rare variants, and *τ*_0_ denoting for the liability threshold of manifesting disease. Here, we assume that a rare variant, PRS and unexplained liability act additively on the liability scale. For simplicity, the PRS *g*_*i*_ is centered and scaled so that *E*[*g*_*i*_] = 0 and 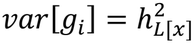 in population, where 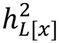 is the heritability explained by PRS on the liability scale. In a case/control cohort,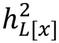 can be inferred from the Area Under the Curve (AUC) using the procedure described in Wray et al.^37^ The mean PRS can be readily estimated from a large unascertained population without using disease outcome data. Under the liability threshold model, the disease threshold *τ*_0_ is determined from the prevalence *K* by Φ^−1^(1 − *K*), where Φ is a standard normal cumulative density function.

Under the above genetic model, we evaluate the likelihood of observed data given the effect size of rare variants *γ* and carrier frequency *f* of the rare variants in population. Specifically, we model the likelihood using the chain rule as follows:

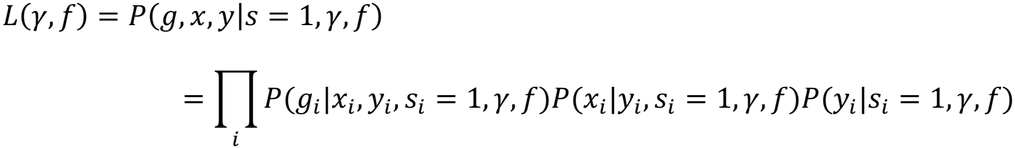

where *s*_*i*_ is an indicator variable denoting whether an individual *i* was sampled into a case/control cohort from population. Note that *s*_*i*_ = 1 for all *i* since all subjects were included in the cohort. *s*_*i*_ is used to account for the case/control ascertainment bias as described later.

The conditional probability of PRS, namely, *P*(*g*_*i*_|*x*_*i*_, *y*_*i*_, *s*_*i*_ = 1, *γ, f*), is assumed independent of *s*_*i*_ or *f* once *x*_*i*_, *y*_*i*_, and *γ* are given. The case/control inclusion or frequency of rare variant carriers are unlikely to affect the distribution of PRS directly. As such, we can simplify *P*(*g*_*i*_|*x*_*i*_, *y*_*i*_, *s*_*i*_ = 1, *γ, f*) with *P*(*g*_*i*_|*x*_*i*_, *y*_*i*_, *γ*), which in turn is modeled using a normal approximation similarly as in Wray et al.^37^ Specifically, for a carrier of rare variants, the conditional probability of PRS among cases and controls follows the following distribution:

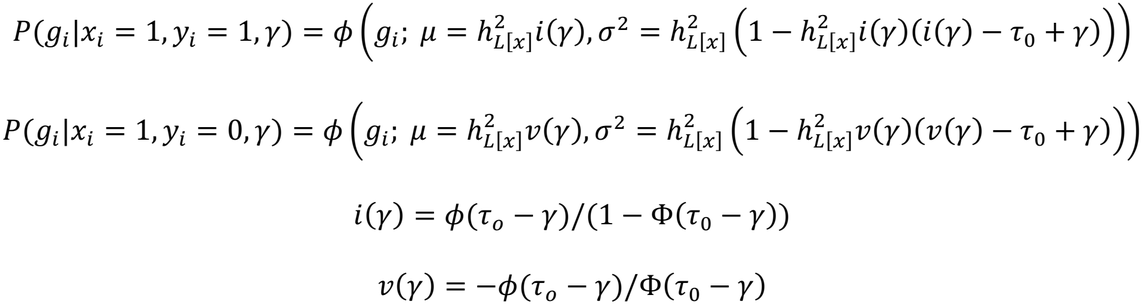

where *ϕ* is a normal probability density function parametrized by *µ* and *σ*^2^. For an individual without rare variants (*x*_*i*_ = 0), the conditional probability of PRS becomes equivalent to the case that *x*_*i*_= 1 with *γ* = 0.

The conditional probability of rare variant genotype, *P*(*x*_*i*_|*y*_*i*_, *s*_*i*_ = 1, *γ, f*), is modeled asbelow:

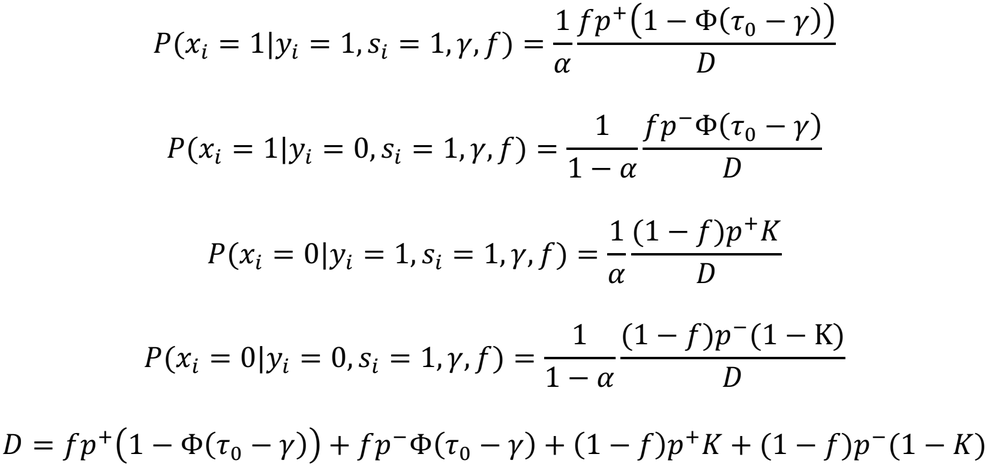

where *α* is the fraction of cases in the cohort *P*(*y*_*i*_ = 1|*s*_*i*_ = 1), *p*^+^ is the likelihood of inclusion as a case if a subject is affected, i.e., 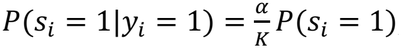, and *p*^−^ is the likelihood of inclusion as a control if a subject is healthy, i.e., 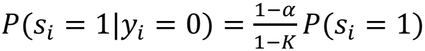. The ascertainment bias in case/control cohorts is accounted for by conditionalizing on *s*_*i*_similarly as in Hosmer and Lemeshow^47^ (See **Appendix A** for the details).

The conditional probability of disease outcome *P*(*y*_*i*_|*s*_*i*_ = 1, *γ, f*) is independent of the effect size of rare variant *γ* and frequency of rare variant carriers *f* given *s*_*i*_ = 1. Thus, we model that:

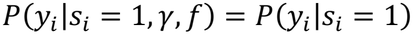

which is *α* if *y*_*i*_= 1 and 1 − *α* if *y*_*i*_ = 0.

The most likely value of *f* can be inferred by the maximum likelihood estimator (MLE) of the full likelihood function as below:

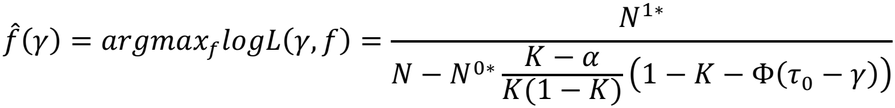

where *N*^1*\**^ is the number of samples with *x*_*i*_ = 1 (rare variant carriers), *N*^0*\**^ is the number of samples with *x*_*i*_ = 0 (noncarriers), and *N* is the total sample size. The estimated value of *f* depends on *γ* since the frequency of carriers increases among cases with a higher *γ* (See **Appendix B** for the details).

The p-value of rare-variant association test is calculated by comparing the log likelihood of observed data under the null hypothesis *H*_*0*_: *γ* = 0 with that under the alternative hypothesis *H*_*1*_: *γ* ≠ 0. Specifically, the likelihood ratio test statistic λ is constructed as follows:

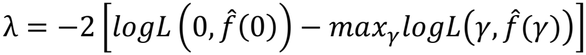

Note that the profile likelihood is used here by replacing *f* with the MLE estimate 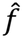. As such, the test statistic λ asymptotically follows a χ^2^ distribution of one degree of freedom under *H*_*0*_. The maximum likelihood is numerically scanned in a grid search over *γ* between *γ* = −5 (near complete protection) to *γ* = +5 (near complete penetrance) in the step size of 0.05.

### Simulated benchmark

To evaluate the Type I error rate of LTBT, 10 million datasets of case/control cohorts were generated for each simulated parameter under the null genetic model with *γ* = 0. A wide spectrum of disease prevalence and predictive power of PRS were tested by exploring *K* = 0.005, 0.01, 0.05 and 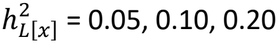. Specifically, for each individual, *g*_*i*_ was randomly sampled from a normal distribution 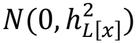, and *ε*_*i*_ was sampled from 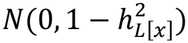.

Genotype *x*_*i*_ was sampled with replacement from the real genotypes of 100,000 White British individuals in the UK Biobank. The real genotype data are based on the rare predicted loss-of-function (pLoF) variants annotated with LOFTEE^42^ in random genes. The rare variants were restricted to the minor allele frequency (MAF) < 0.01, and the random genes was selected from 1,974 genes with the cumulative allele frequency (CAF) of rare variants > 0.0005. The case/control cohorts were generated by simulating an unascertained population of 2,000,000 subjects and then randomly down-sampling to the target case and control counts.

To compare the statistical power of LTBT with existing methods, 1 million benchmark datasets were generated for each *γ* from 0.2 to 1.0, similarly as above. All rare variants were assumed causal with the same direction and effect size (*γ*). To account for multiple testing of 50,000, the *α* level was set to 10^−6^. The case/control cohort was generated for the target sample size of N = 5,000 cases and 5,000 controls. The prevalence was varied over *K* = 0.005, 0.01, and 0.05. The prediction accuracy of PRS was explored over 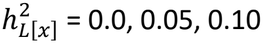, and 0.20.

For the comparison with LTBT, a burden test, fisher’s exact test, and SKAT-O^38^ were benchmarked on the same simulated datasets. A burden test was done under a logistic regression model with collapsed genotypes in a dominant model. Fisher’s exact test was similarly performed under a dominant genetic model. SKAT-O was run in a default setting using SKATBinary_Robust function in R using saddle point approximation.^48^

### Discovery cohort

For a discovery cohort, 136 pulmonary fibrosis patients of European ancestry from the Clinical Genetics and Screening for Pulmonary Fibrosis (CGS-PF) study were combined with ancestry-matched controls from the UK Biobank.^49^ The CGS-PF cohort is a family-based cohort established to determine merits of screening first-degree relatives of patients with IPF for evidence of early disease.^40^ While the CGS-PF includes both probands and at-risk relatives, the current study utilized only probands for our discovery analysis. The inclusion criterion for probands in CGS-PF was to be aged 48-85 years with chest CT evidence of pulmonary fibrosis (defined by pulmonary parenchymal architectural distortion) and clinically evident, or radiologically suspected. The diagnosis is determined by the referring pulmonary physician using standard criteria defined by the American Thoracic Society. Genomic DNA was collected from peripheral blood samples using PAXgene Blood DNA tubes. Whole-Exome Sequencing (WES) was performed in a CLIA-certified lab at the Yale Center for Genome Analysis. Exome capture was conducted with IDT xGen Exome Hyb Panel targeting 205,398 regions (a total of 39.4 Mbp). The capture products were pooled and sequenced in the NovaSeq S4 flowcell for 100 bp paired-end sequencing. This study was conducted under the approval of Institutional Review Board of Mass General Brigham (#2016P000837).

To make sure that probands are all unrelated, their familial relationship was examined across CGS-PF samples by running KING^50^ on genetic data. In addition, to select probands with European ancestry, CGS-PF samples were projected into the principal component analysis (PCA) space constructed with genetic data of reference population panels from the 1000 Genomes Project (phase 3 v5). For both familial relationship and PCA analyses, WES data were used as genetic data. Variants were filtered, keeping only those with MAF > 0.05 and missing rate < 0.05, and LD-pruned at the window size of 50 kb, step size of 2, and r^2^ threshold of 0.2.

For controls, a total of 230,496 White British individuals were selected from the UK Biobank as described below. First, using the death registry and in-patient diagnosis, all subjects diagnosed with any history of interstitial lung disease (ILD, ICD10: J84) were filtered out from the control. The filtering by ILD rather than narrower definition of IPF was intentional to account for the possibility of underdiagnosis of IPF in the UK Biobank. Second, all participants with a self-reported history of ILD (asbestosis, pulmonary fibrosis, and fibrosing alveolitis) were also excluded from controls based on questionnaire data. Third, individuals withdrawn from the study, aged less than 65, of non-European ancestry, or flagged with any QC issues were filtered out. The QC issues include the sex mismatch, sex chromosome aneuploidy, outlier for the heterozygosity or missing rate, or exclusion from kinship inference. Lastly, only unrelated subjects were retained as controls by keeping only one sample for each family based on familial relationship data.

To minimize potential stratification between cases and controls, a uniform variant calling and QC pipeline was performed on WES data from CGS-PF and UK Biobank. First, the variant calling pipeline based on DeepVariant^51^ and GLNexus^52^ was replicated on CGS-PF data under the identical parameter setting as they were used in the UK Biobank project.^8^ Second, after the variant calling, the same variant QC criteria were applied to both datasets. Variants are retained only if the read depth (DP) is ≥ 10 for at least 90% of samples, the call rate is > 0.9, and the Hardy-Weinberg equilibrium p-value > 10^−15^, and the alleles are well balanced. The allele balance (AB) of alternative allele is required to be < 0.1 for reference homozygotes, > 0.9 for alternative homozygotes, and between 0.2-0.8 for heterozygotes. Variants that failed to satisfy the QC criteria in either CGS-PF or UK Biobank were excluded from both datasets.

Then, once cases and controls were combined into one cohort, all common variants with the MAF > 0.001 or POPMAX > 0.01 in gnomAD (release 2.1.1)^43^ were filtered out. For the functional annotation of variants, Ensembl VEP (version 111) was used.^53^ Computationally predicted loss-of-function (pLoF) variants were identified using Loftee.^42^ Probably damaging missense variants were identified using PolyPhen 2.^43^

### Validation cohort

All of Us Biobank (CDR v8)^44^ was used to formally replicate the association signals found in the discovery analysis. The case subjects were selected by taking the subset of individuals diagnosed with IPF (ICD10: J84.1) and taking antifibrotic medications, specifically, nintedanib (concept id: 45775206) and pirfenidone (concept id: 45775396) in electronic health records.

The control subjects were all individuals with ages of 65 or higher and no history of IPF. Both cases and controls were restricted to the individuals of European genetic ancestry. Samples flagged with QC issues were excluded. Related samples were filtered out based on genetic relatedness data.

## Data Availability

All data produced in the present study are available upon reasonable request to the authors. Software developed for this study is available online at https://github.com/sgchun/ltbt.

## SOFTWARE AVAILABILITY

The source codes for LTBT are available at https://github.com/sgchun/ltbt along with QC scripts for the UK Biobank and All of Us Biobank.

## ACKNOWLEDGMENTS

S.C. was supported in part by NIH grant R56HL171620. This research has been conducted using the UK Biobank Resource under Application Number 62142. This research has been conducted also using the All of Us Research Program data and would not be possible without the contributions of individuals who take part in the All of Us Research Program.

G.M.H. and this work was supported by RO1s HL111024, HL135142, and HL130974.

## Appendix A. Conditional probability of rare-variant genotype given disease outcome.

We account for the ascertainment bias in case/control cohorts by conditionalizing on the sample inclusion selection variable *s*_*i*_ = 1 similarly using the strategy of <u>Hosmer and Lemeshow</u> as follows. By Bayes theorem,

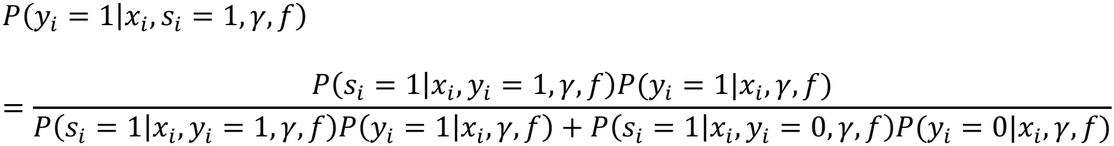

and

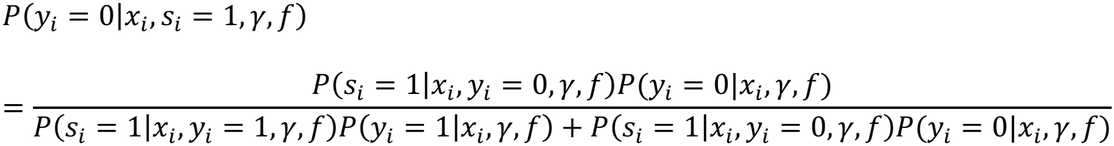

Under the assumption that the sample selection *s*_*i*_ depends only on disease status *y*_*i*_, *P*(*s*_*i*_ = 1|*y*_*i*_, *x*_*i*_, *γ, f*) = *P*(*s*_*i*_ = 1|*y*_*i*_). Let us put *p*^+^ = *P*(*s*_*i*_ = 1|*y*_*i*_ = 1) and *p*^−^ = *P*(*s*_*i*_ = 1|*y*_*i*_= 0). Then,

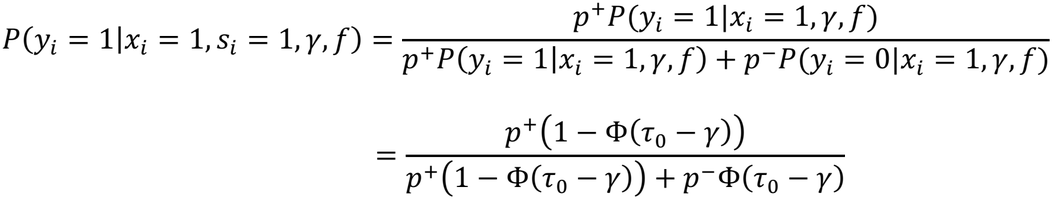

since *P*(*y*_*i*_ = 1|*x*_*i*_ = 1, *γ, f*) = 1 − Φ(*τ*_0_ − *γ*) and *P*(*y*_*i*_ = 0|*x*_*i*_ = 1, *γ, f*) = Φ(*τ*_0_ − *γ*). Similarly,

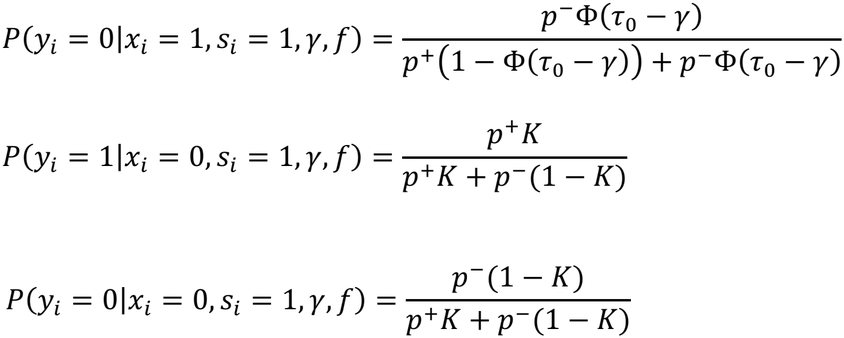

since *P*(*y*_*i*_ = 1|*x*_*i*_ = 0, *γ, f*) = 1 − Φ(*τ*_0_) = *K* and *P*(*y*_*i*_ = 0|*x*_*i*_ = 0, *γ, f*) = Φ(*τ*_0_) = 1 − *K*.

Then, *P*(*x*_*i*_= 1, *s*_*i*_ = 1|*γ, f*)

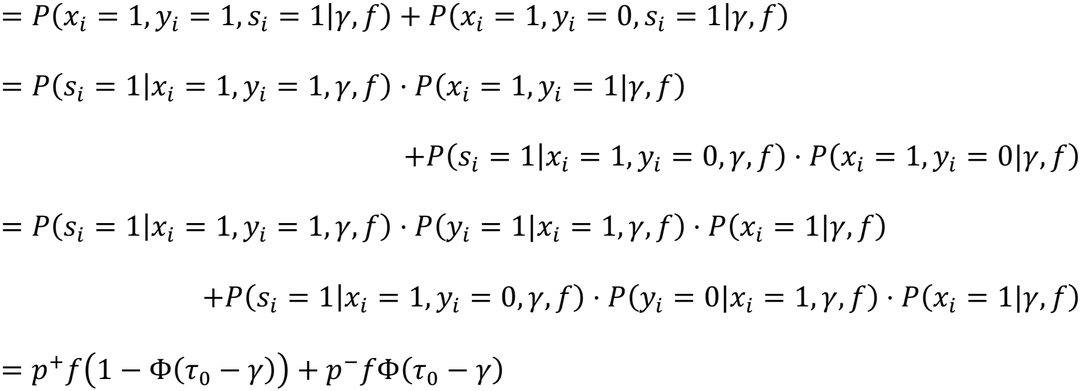

since *P*(*x*_*i*_ = 1|*γ, f*) = *f*. Similarly, *P*(*x*_*i*_ = 0, *s*_*i*_= 1|*γ, f*)

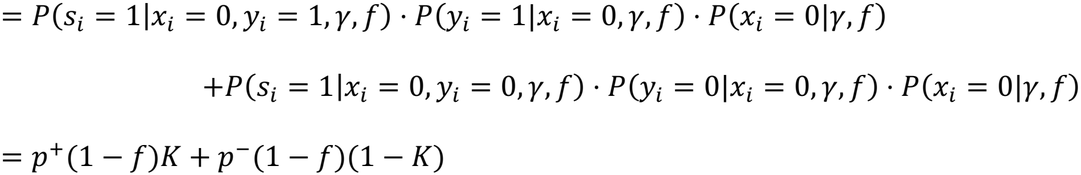

By conditional probability,

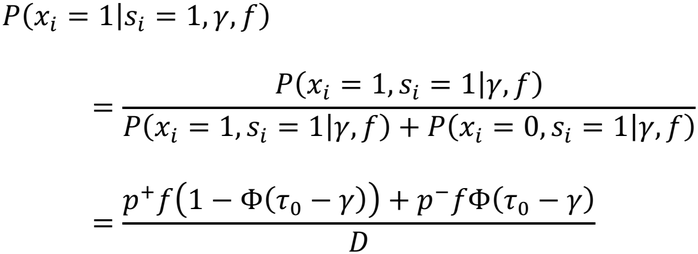

where *D* = *p*^+^*f*01 − Φ(*τ*_0_ − *γ*)4 + *p*^−^*f*Φ(*τ*_0_ − *γ*) + *p*^+^(1 − *f*)*K* + *p*^−^(1 − *f*)(1 − *K*). Similarly,

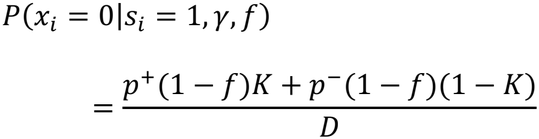

Therefore, from the above derivations,

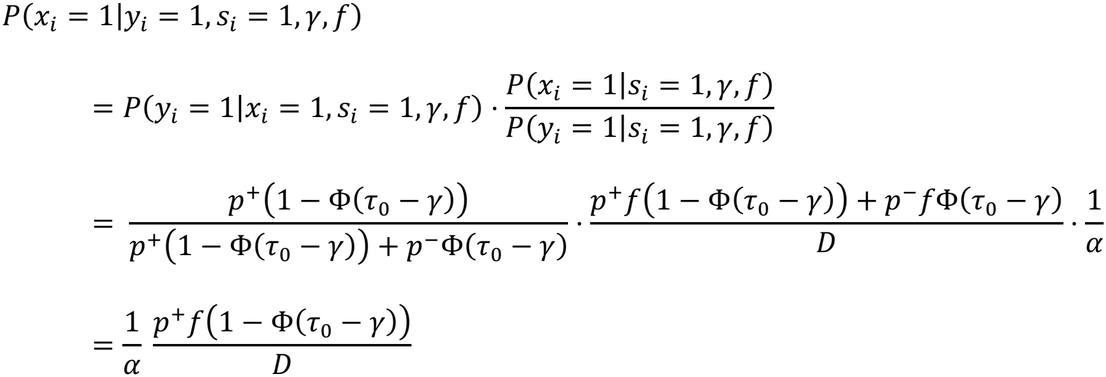

Similarly,

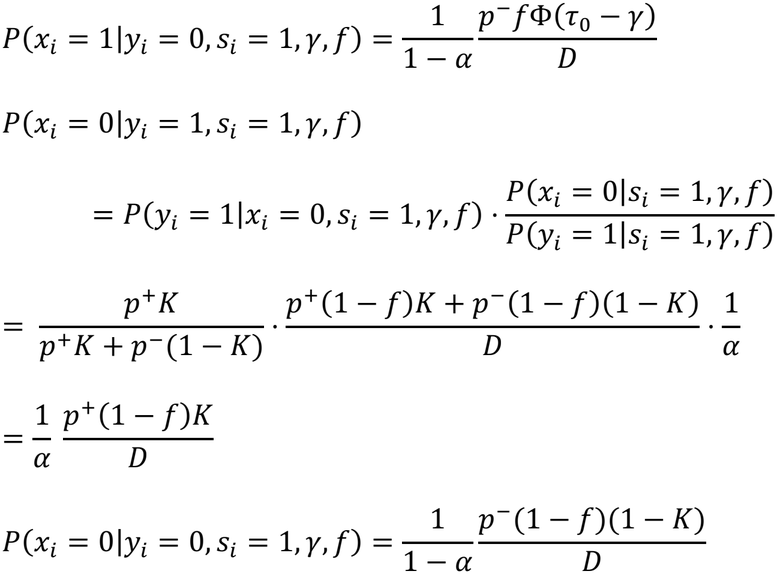

## Appendix B. Maximum likelihood estimator of the carrier frequency of rare variants *f*.

Given a fixed *γ*,

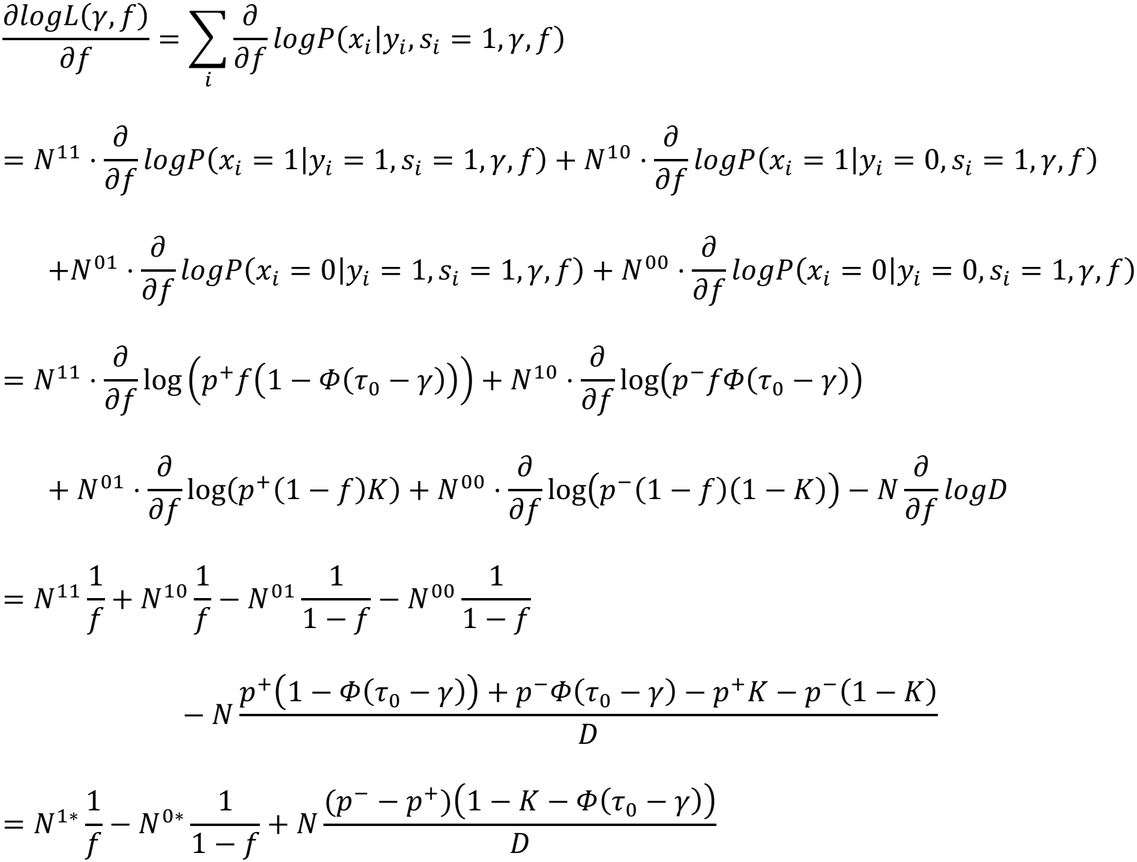

where *N*^11^ is the number of case samples with *x*_*i*_ = 1, *N*^10^ is the number of control samples with *x*_*i*_ = 1, *N*^01^ is the number of case samples with *x*_*i*_ = 0, *N*^00^ is the number of control samples with *x*_*i*_ = 0, *N*^01^ is the number of samples with *x*_*i*_ = 1, *N*^00^ is the number of samples with *x*_*i*_ = 0, and *N* is the total sample size. Since *D* simplies to:

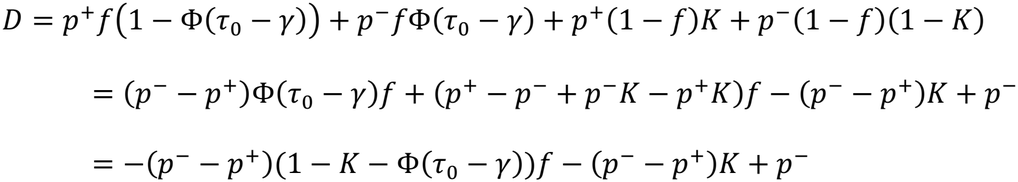

The maximum likelihood estimator for *f* given a fixed *γ* can be analytically derived by solving the following for *f*:

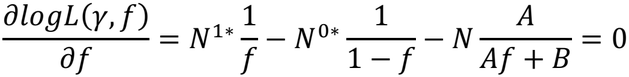

where *A* = (*p*^*#x2212;^ − *p*^+^)01 − *K* − *Φ*(*τ*_0_ − *γ*)4 and *B* = (*p*^−^ − *p*^+^)*K* – *p*^−^. By simplifying the equation,

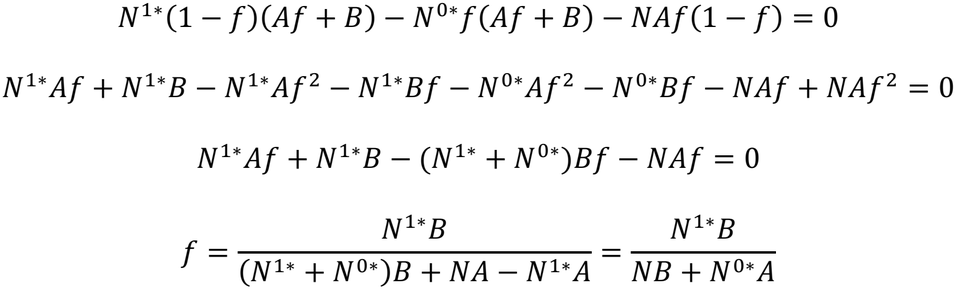

since *N*^1*\**^ + *N*^0*\**^ = *N*.

From 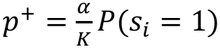, and 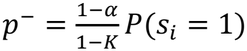,

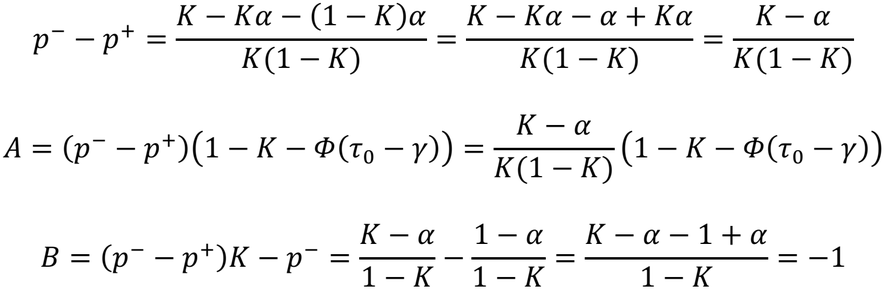

Thus, in general, the maximum likelihood estimator of *f* becomes:

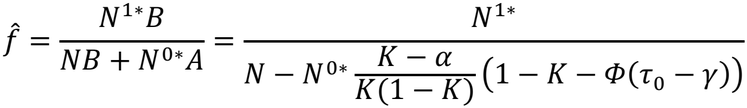

In a special case of unascertained population cohort, *α* = *P*(*y*_*i*_ = 1|*s*_*i*_ = 1) = *K*, thus

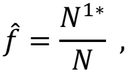

which is equivalent to the sample frequency of rare-variant carriers in the cohort. In addition, when *γ* = 0 under the null hypothesis, *Φ*(*τ*_0_ − *γ*) = 1 − *K*, and 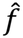 also simplifies to:

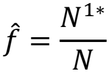

## Notes

### Competing Interest Statement

The authors have declared no competing interest.

### Author Declarations

The IRB of Mass General Brigham gave approval for this work (#2016P000837).

